# The effect of ABO blood group and antibody class on the risk of COVID-19 infection and severity of clinical outcomes

**DOI:** 10.1101/2020.09.22.20199422

**Authors:** Marwa Ali Almadhi, Abdulkarim Abdulrahman, Abdulla Alawadhi, Ali A.Rabaan, Manaf AlQahtani

## Abstract

The current COVID-19 pandemic has affected more than 22 million cases and caused immense burdens on governments and healthcare systems worldwide. Since its emergence in December 2019, research has been focused on ways to not only treat the infected but also identify those at risk and prevent spread. There is currently no known biological biomarker that can predict the risk of being infected. A growing set of studies have emerged that show an association between ABO blood group and the risk of COVID-19 infection. In this study, we used retrospective observational data in Bahrain to investigate the association between ABO blood group and risk of infection as well as susceptibility to a more severe ICU-requiring infection. We found that individuals with blood group B were at a higher risk of infection, while those with blood group AB were at a lower risk. No association was observed between blood group and the risk of a severe ICU-requiring COVID-19 infection. We extended the analysis to study the association by antibodies present; anti-a (blood groups B and O) and anti-b (blood groups A and O). Antibodies were not found to be associated with either risk of infection or susceptibility to severe infection. The current study, along with the variation in blood group association results, indicates that blood group may not be the most ideal biomarker to predict risk of COVID-19 infection.

## BACKGROUND AND SIGNIFICANCE

Severe Acute Respiratory Syndrome Coronavirus 2 (SARS-CoV-2), the virus causing the current COVID-19 pandemic, has led to over 22 million cases and 788,000 deaths worldwide (1). The rapid spread of the disease has inflicted immense strains on healthcare and testing resources, and identifying individuals most at risk is critical to managing the pandemic. There is currently no known biological biomarker that can predict the risk of being infected. Several studies have emerged showing an association between ABO blood types and the risk of COVID-19 infection. This is not unlikely, as many studies previously suggested associations between blood group and other diseases and infections, including SARS-CoV-1 (2–4). This observation was first reported by Zhao *et al*. for SARS-Cov-2, with blood group A showing a higher risk of COVID-19 infection and mortality, and blood group O showing a decreased risk (5). These findings have been replicated in several other reports (6–9). Further analysis of this data was conducted by antibodies, classifying blood groups as anti-A (blood groups B and O) and anti-B (blood groups A and O), and suggested that anti-A antibodies are less associated with COVID-19 (10). However, a recent article by Latz *et al*. reports contradicting observations, where higher risk of infection was observed for individuals of blood group B instead of A (11). This recent contradiction to the literature has added ambiguity to the field. Hence, our objective is to identify whether the risk of COVID-19 infection and severity of clinical outcomes are associated with ABO blood groups and antibodies.

## METHODS

### 1. Study Design

In this cross-sectional observational study, we investigated the association between blood group and risk of COVID-19 infection and severity of clinical outcomes. Association was analyzed by ABO blood group and blood antibody class. To study effect of blood group on susceptibility of a SARS-CoV-2 infection, the distribution of blood types amongst a sample of confirmed COVID-19 individuals in Bahrain were compared to the distribution of blood types amongst the general population of Bahrain. To study the effect of blood type on the severity of clinical outcomes, the distribution of blood types were compared between COVID-19 individuals who were admitted to an intensive care unit (ICU) and those of the general COVID-19 patients sample. This was replicated in the analysis by antibody class, with blood groups being grouped as anti-A (blood groups B and O), and anti-B (blood groups A and O).

### 2. Data collection

A confirmed COVID-19 case was any individual who tested positive for SARS-CoV-2 via a nasopharyngeal (NP) swab. Presence of SARS-CoV-2 in the NP sample was tested by polymerase chain reaction (PCR) analysis using the E gene as a target. If the E gene was detected, the sample was confirmed by a PCR test targeting RdRp. All confirmed cases tested positive for the E gene and the RdRp using real time PCR from Roche and Invitrogen kits.

A random sample of 3000 COVID-19 positive individuals was chosen from the National COVID-19 Database to represent the COVID-19 infected population. Of those, 2138 individuals had blood group data documented or obtainable, and hence included in the study. As of 19 July 2020, there have been 196 COVID-19 cases with ICU admissions, all of which were included in this study. Blood group data for all the COVID-19 infected individuals were obtained from medical records and the National COVID-19 Database. To represent the blood type distribution of the general population of Bahrain, the blood types of 4985 individuals who donated whole blood or platelets at King Hamad University Hospital Blood Bank over the past 2 years were obtained and analyzed.

### 3. Statistical analysis

Chi-squared (X^2^) and Fischer’s exact tests were used to compare the distributions of blood groups and antibodies between samples. Odds Ratio (OR) tests were used to study the odds of a blood type or antibody category testing positive, in a one-vs-all manner. ORs are reported with 95% confidence intervals. Statistical values were considered significant at p<0.05. Statistical analysis was performed using STATA statistical software (version 15.1).

## RESULTS

### 1. Analysis of susceptibility to COVID-19 infection

Of 4985 individuals representing the control group, we observed a high frequency of blood group O (n=2316, 46.46%), followed by B (n=1225, 24.57%), A (n=1092, 21.91%) and AB (n=352, 7.06%) accordingly. A total of 2334 COVID-19 infected individuals represented the COV+ group, showing an identical order of frequency; blood group O (n=1060, 45.41%), followed by B (n=644, 27.59%), A (n=513, 21.98%) and AB (n=117, 5.01%). The distributions of the two groups are shown in **Figure 1**. Blood group distributions were statistically different between the two groups (X^2^ = 16.45, p = 0.001). Logistic regression analysis showed that blood group AB was associated with a decreased risk of infection (OR: 0.69, 95% CI: 0.56 – 0.86, p=0.001) while blood group B was associated with an increased risk (OR: 1.17, 95% CI: 1.04 – 1.31, p=0.006). No association between blood group A and risk of COVID-19 infection was found (OR: 1.00, 95% CI: 0.89-1.13, p = 0.943)(**Table 1**). Analysis by antibody class showed no association between type of antibodies present and risk of testing positive (**Table 1**).

**Table 1.** Comparison of ABO blood group distributions and antigens present between the general population and COVID-infected sample.

| Blood type | X<sup>2</sup> | OR | 95% CI | P-value |
| --- | --- | --- | --- | --- |
| A | 0.28 | 1.10 | 0.76 – 1.56 | 0.599 |
| B | 0.67 | 1.14 | 0.82 – 1.59 | 0.411 |
| O | 1.83 | 0.81 | 0.60 – 1.11 | 0.177 |
| AB | 0.16 | 1.14 | 0.54 – 2.17 | 0.688 |
| Anti-A | 0.47 | 0.89 | 0.64 – 1.26 | 0.491 |
| Anti-B | 0.94 | 0.86 | 0.63 – 1.18 | 0.332 |

**Figure 1.**
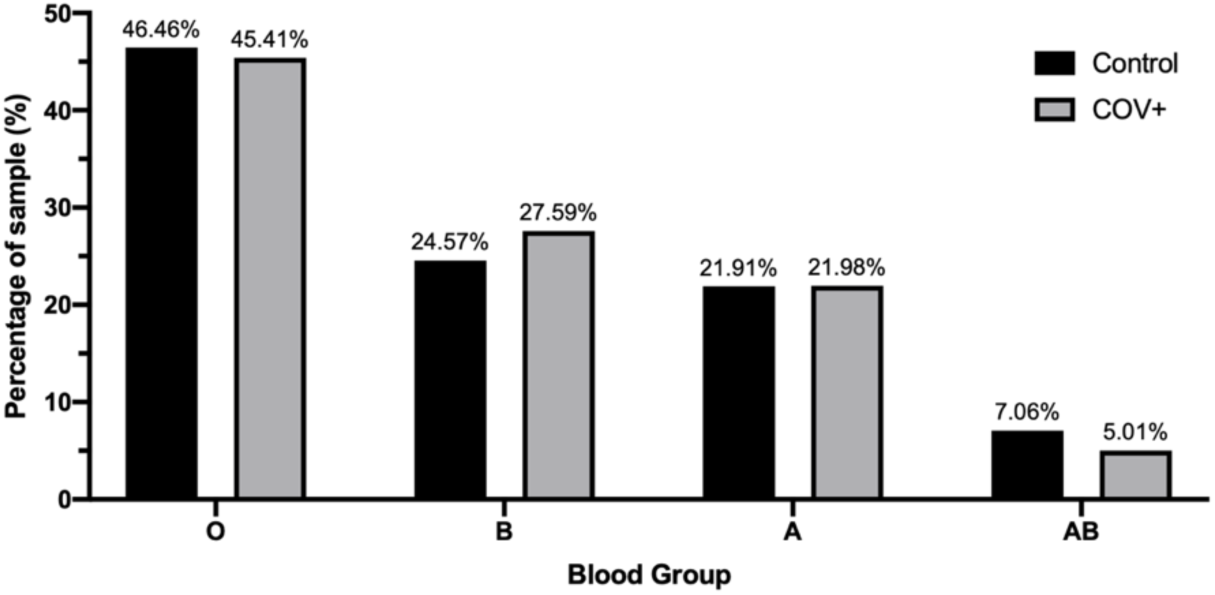
Blood group distributions in the non-COVID-19 (control) and COVID-19 positive (COV+) samples.

### 2. Analysis of severity of COVID-19 infection

To test the association between blood group and the susceptibility to a severe COVID-19 infection, blood group distributions were compared between COVID-19 infected individuals that required ICU admission and those that did not, COV+ICU+ and COV+ICU-respectively. Of 196 COV+ICU+ individuals, 80 (40.82%) were blood group O, 59 (30.10%) were blood group B, 46 (23.47%) were of blood group A, and 11 (5.61%) were of blood group AB. Of 2138 COV+ICU-individuals, 980 (45.84%) were blood group O, 585 (27.36%) were blood group B, 467 (21.84%) were of blood group A, and 106 (4.96%) were of blood group AB. The distributions of the two groups are shown in **Figure 2**. No difference in blood group distributions was observed (X^2^ = 1.85, p = 0.603). No association to severity of COVID-19 infection was found with blood group or antibodies present (**Table 2**).

**Table 2.** Comparison of ABO blood group distributions and antigens present between COV+ICU- and COV+ICU+

| Blood type | X<sup>2</sup> | OR | 95% CI | P-value |
| --- | --- | --- | --- | --- |
| A | 0.01 | 1.00 | 0.89 – 1.13 | 0.943 |
| B | 7.62 | 1.17 | 1.04 – 1.31 | 0.006 |
| O | 0.70 | 0.96 | 0.87 – 1.06 | 0.404 |
| AB | 11.12 | 0.69 | 0.56 – 0.86 | 0.001 |
| Anti-A | 3.05 | 1.10 | 0.99 – 1.23 | 0.081 |
| Anti-B | 0.69 | 0.96 | 0.86 – 1.06 | 0.407 |

**Figure 2.**
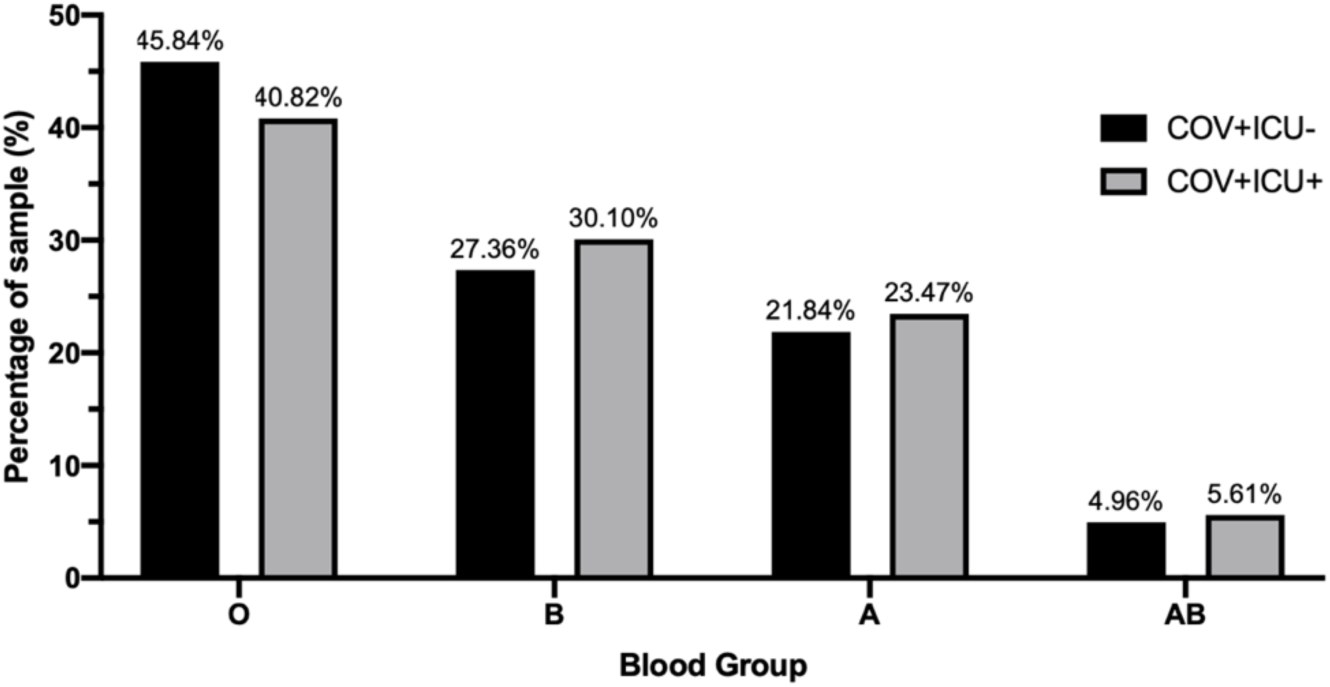
Blood group distributions in non-ICU (COV+ICU-) and ICU (COV+ICU+) COVID-19 positive samples.

## DISCUSSION

Following several reports of the association between blood group distribution and risk of COVID-19 or susceptibility to severe infection, we sought to analyse these trends amongst the population in Bahrain. This study used blood-bank data of blood distributions to represent the general population in Bahrain. The distributions we obtained were similar to a previous report of blood-bank blood distributions, which also showed that these distributions were comparable to population sample distributions in Bahrain (12).

Regarding the association between blood group and risk of COVID-19 infection, most studies were similar in reporting that blood group A was associated with an increased frequency amongst COVID-19 individuals and risk of infection, and conversely that blood group O was associated with a decreased frequency and risk of infection (5–9). Our data does not present an association between susceptibility to COVID-19 infection and blood group A, similar to observations by Latz *et al*. (11). In addition, we found a significantly increased risk associated with blood group B, which is also comparable to findings by Latz *et al*. (11). Individuals with blood group AB showed a decreased risk of COVID-19 infection, synonymous with a few reports, however in all these cases including this study, this group was represented by a small sample size (5, 6). Unlike all other reports, no association between blood group O and decreased risk of infection was observed.

To study the association of blood group and the severity of COVID-19 infection, we compared blood group distributions between the general COVID-19 sample and those that required ICU admission. As previously reported, no association was observed between blood group and susceptibility to a more severe COVID-19 infection (6–8, 11). Although there was a decrease in odds of individuals with blood group O requiring ICU admission, similar to observations by Zhao *et al*., this was not statistically significant (5). Hence, no association was observed between blood group and susceptibility to a more severe infection.

As per Gerard *et al*., we extended the analysis by antibody class (10). There was no association found between antibodies present and the risk of COVID-19. We observed that individuals with anti-A antibodies were at higher odds of testing positive for COVID-19, however this finding was not statistically significant. This finding contradicts that by Gerard et al, who found that anti-A was associated with a significantly lower risk of infection. There is no published analysis yet of the relationship between antibodies and severity of outcome, however in this study we found none.

Results from this study, and the limited reports in this field, reveal a variety of findings making a conclusion regarding an association between blood type and COVID-19 challenging. However, this variation in results may indicate that an unexplored underlying factor may be causing the association, not necessarily the blood group or type of antibodies present.

## Data Availability

The data that support the findings of this study are available on request from the corresponding author.

## Declaration

### Ethics approval

Approval received from National COVID-19 Research Committee

### Consent to participate

Not applicable

### Consent for publication

All author approved to publish this data

### Availability of data and material

Available

### Code availability

All data were entered in Microsoft excel and Stata

### Authors’ contributions

Manaf AlQahtani contributed to the study conception and data collection. AbdulKarim AbdulRahman contributed to the study design, data preparation and study supervision Abdulla AlAwadhi contributed in Material preparation and data collection. design. Marwa AlMadhi contributed to the data analysis and wrote the first draft of the manuscript. Ali Rabaan reviewed the manuscript All authors commented on previous versions of the manuscript. All authors read and approved the final manuscript.

